# Diagnostic Classification for Long Covid Patients identifying Persistent Virus and Hyperimmune Pathophysiologies

**DOI:** 10.64898/2026.04.21.26351402

**Authors:** Philip H. James-Pemberton, Daniel Harper, Poppy J. Wagerfield, Ciara E. Watson, Luca Hervada, Shivali Kohli, Zachary J. Rees, Ruju Mistry, Steve Allder, Andrew M. Shaw

**Affiliations:** Biosciences, University of Exeter, Stocker Road, Exeter, EX4 4QD, UK; Attomarker Ltd, 3 Babbage Way, Exeter Science Park, Exeter, Devon, EX5 2FN, UK; Re:Cognition Health Clinic in the UK · London. 77 Wimpole Street London, W1G 9RU

## Abstract

A multiplex diagnostic test is evaluated for self-reported long COVID associated persistent symptoms and a poor recovery from a SARS-CoV-2 infection. A mass-standardised concentration of total antibodies (AC), high-quality (HQ) antibodies and percentage of HQ antibodies (HQ%) is assessed against a spectrum of spike proteins to the SARS-CoV-2 variants: Wuhan, α, δ, and the Omicron variants BA.1, BA.2, BA.2.12.1, BA.2.75, BA.5, CH.1.1, BQ.1.1 and XBB.1.5 in three cohorts. A cohort of control patients (*n* = 46) recovered (CC) and a cohort of self-declared long COVID patients (*n* = 113) (LCC). A nested Receiver Operating Characteristic (ROC) analysis, performed for the variant with lowest HQ concentration in the spectrum, produced an area under the curve and AUC = 0.61 (0.53-0.70) for the CC *vs* LCC cohorts. For the LCC cohort, the cut-off thresholds for AC = 0.8 mg/L, HQ = 1.5 mg/L and HQ% of 34% were determined, leading to a 71% sensitivity and 66% specificity derived by the Youden metric. The cohorts may be fully classified based on ROC and outlier analysis to give an incidence of persistent virus 62% (95% CI 52% – 71%), hyperimmune 12% (95% CI 7% - 20%) and unclassified, 26% (95% CI 18% - 35%). The overall diagnostic accuracy for both the hyper and hypo immune is 69%. All clinical interventions can now be tailored for the heterogenous long COVID patient cohort.

## Introduction

Post-viral sequalae following a SARS-CoV-2 infection have been characterised as a syndrome of more than 250 symptoms called long COVID^1^. WHO has attempted to standardise the definition of long COVID^2^ based on the presence of a selection of symptoms more than three months after the initial infection, lasting at least two months, and with no other explanation. Prevalence of long COVID globally has now been estimated at 400 million^3^; in the US prevalence for adults in 2024 was reported as 6.9%, or 17m people^4^. An accurate diagnostic test would be the beginning of a coherent response and treatment for patient but a conventional diagnostic test analysis, such as a Receiver Operator Characteristic (ROC) requires a well-defined dichotomous marker which is not possible in this cohort. Consequently, the definition of a health or control cohort is also compromised.

The pathophysiology for long COVID is not yet determined although the literature is dominated by two hypotheses: viral persistence and autoimmunity^5-7^. The persistent virus mechanism suggests micro-infection and micro-inflammation events around the body associated with ACE2 receptors^8^ (heart, lungs, brain and gut, for example) which would explain the frequently reported symptoms such as microbiome dysbiosis, postural orthostatic tachycardia syndrome (POTS) and brain fog, amongst others^1^. Autoimmunity could affect any tissue based on random association of an antibody raised to SARS-CoV-2 and a corresponding epitope on a tissue protein. Autoantibodies to the ACE2 receptor, β - adrenoreceptor, muscarinic M2 receptor and angiotensin II AT receptor have been reported^9^.

Many studies have sought a biomarker to classify long COVID patients as having an uncoordinated T and B cell response^10^; microclots resistant to fibrinolysis^10^, high levels of autoantibodies that were inversely correlated with neutralising SARS-CoV-2 antibodies^10,11^; increased levels of antibodies against the SARS-CoV-2 spike and the S1 subunit compared to vaccinated individuals^10^; and sustained levels of spike protein over the course of several months. There was also circulating spike protein detected in 60% of the long COVID patient samples tested^12^. None appear to have performed a ROC analysis.

We hypothesise that a persistent viral reservoir would arise from a poor quality non-sterilising serum consisting of a low concentration of poor-quality antibodies. In this study, we characterise the antibody profile to a spectrum of variant spike proteins to SARS-CoV-2 measuring total antibody concentration, AC, raised to a panel of 11 variants, and the concentration of high-quality antibodies, HQ, that survive binding to the spike protein of each variant at pH 3.2 (HQ) correlating to a long paratope-epitope complex half-life required for efficient removal of the virus. The percentage of high-quality antibodies HQ% = HQ/AC × 100 is also calculated and is independent of the HQ, AC concentrations. These three measures were derived for a spectrum of variant spike proteins: Wuhan, α, δ, and the Omicron variants BA.1, BA.2, BA.2.12.1, BA.2.75, BA.5, CH.1.1, BQ.1.1 and XBB.1.5. Two patient cohorts were tested: a control cohort collected (*n* = 46) at by Attomarker (CC) and a commercially collected cohort of patients self-referring with long COVID through private clinics (LCC) (*n* = 113), taking the test on a named-patient basis under the recommendation of the clinician.

The persistent virus long COVID risk was identified as the lowest HQ antibody levels of any variant, defining an AC, HQ and HQ% triplet for each patient. A triple-nested ROC analysis was performed for the CC and LCC cohorts against the self-reporting dichotomous long COVID marker based on symptoms. A cut-off for each parameter was determined using the Youden Index maximum and the Euclidean metric. The ROC thresholds are considered as characteristic properties of a sterilising serum. A distribution outlier analysis is considered as a diagnostic classification for hyperimmune patients which is combined with the ROC analysis to classify all long COVID patients suggesting a hypo-immune and hyper-immune classification indicated by the two proposed pathophysiologies.

## Materials and Methods

### Biophotonic Multiplexed Immuno-Kinetic assay

The multiplexed biophotonic array platform technology has been described in detail elsewhere^13,14^ and is CE marked to perform the Attomarker COVID Antibody Immunity Test. The current protocol now has a UKCA mark as the COVID Antibody Spectrum Test. Briefly, the technology is a localised particle plasmon gold sensor which scatters light proportional to the mass of material (formally refractive index) near to the gold nanoparticle surface in the plasmon field. The gold nanoparticles are printed into an array of 210 spots which are then individually functionalised with the protein required. The assay consists of a capture step, a detection step and a regeneration step with intervening wash steps. High and low controls are used to calibrate the sensor response and are repeated at regular intervals to ensure quality control^15^ during the day.

Two different sensor arrays were fabricated each containing six spike variant proteins, anti-C-reactive protein, recombinant HSA and protein G. One of the spike variant proteins was common (Wuhan) to allow the results from the two arrays to be compared. The proteins on chip one were: Wuhan (ACROBiosystems, SPN-C82E9), Alpha (ACROBiosystems, SPN-C82E5), Delta (ACROBiosystems, SPN-C82Ec), BA.1 (Sinobiological, 40589-V49H3-B), BA.2 (ACROBiosystems, SPN-C82Er) and BA.5 (Sinobiological, 40589-V49H5-B). The proteins on the second chip were: Wuhan (ACROBiosystems, SPN-C82E9), BA.2.12.1 (Sinobiological, 40589-V49H7-B), BA.2.75 (Sinobiological, 40589-V49H6-B), CH.1.1 (ACROBiosystems, SPN-C82Q0), BQ.1.1 (ACROBiosystems, SPN-C82Ey) and XBB.1.5 (ACROBiosystems, SPN-C82Ez). The common non-spike proteins to each chip were: anti-CRP (Bethyl, A80-125B), recombinant HSA (Sigma, A9731), protein G (ThermoFisher 29988).

The integrity of the spike protein samples on the surface was tested using an anti-S2 antibody (SinoBiological, 40590-D001), chosen for its specificity to a pan-variant conserved epitope. This allows for it to be used in calibrating all spike protein channels simultaneously. The concentration of the antibody was calibrated against the NIST antibody RM8671, NISTmAb, a recombinant humanized IgG1κ with a known sequence^15^ to assure monomeric purity of the antibody. High-control and low-control samples were made using the 40590-D001 antibody and these controls were then used to quantify results from human samples in units of mg/L.

Thus, the assay response is presented as a concentration of an antibody equivalent in its binding properties to the 40590-D001 control antibody.

### Patient Cohorts

Two cohorts were collected; a control cohort of healthy volunteers (CC) and cohort of patients were referred for the research test individually under a named-patient basis through private clinics (in the UK and EU): the LCC cohort. The demographics of the CC and LCC cohorts are detailed in Table 1; the LCC cohort was self-reporting and reported to the study based on a variety of symptoms. The LCC patients were referred for the research test following a consultation and a decision from the clinician that the test was in the patient’s interest.

**Table 1.** Patient cohort demographics.

| **Demographic** | **Controls (CC)** | **Long COVID Clinics (LCC)** |
| --- | --- | --- |
| Number of Patients | 46 | 113 |
| Full Demographics Recorded | 91% | 77% |
| Male/Female | 14/30 | 43/42 |
| Age Range | 18-75 | 14-74 |
| Double vaccinated | 93% | 86% |
| 1<sup>st</sup> Booster Vaccinated | 78% | 70% |
| 2<sup>nd</sup> Booster Vaccinated | 17% | 19% |
| Median (IQR) Time: First Known Infection to Sample (days) | 301 (96-835) | 887 (432-1167) |
| Median (IQR) Date of Collection | 4-Jun-24<br>(13-Jul-22 to 2-Jul-24) | 10-Jun-24<br>(13-Jul-22 to 1-Jul-24) |

### Ethical Approval

The use of the Attomarker clinical samples was approved by the Bioscience Research Ethics Committee, University of Exeter. The LCC cohort was collected under named-patient ethics by the clinicians for the test; the use of the results is approved as above.

### Numerical Analysis

The AC distribution for the CC cohort was analysed for each of the variants to identify hyperimmune outliers using the *Z* score > 2 metric, with the mean value for the three early variants taken as these were significantly higher than the responses to the later variants, Table S2. Samples excluded by this test were not taken forward into the ROC analysis. A ROC analysis was then performed using the dichotomous marker based on the CC cohort and self-reported LCC cohort. The persistent variant risk for each patient was identified by selecting the lowest HQ concentration antibody in the variant spectrum, the variant for which there is least likely to be a sterilising serum. The variables AC, HQ and HQ% were identified for the variant with the lowest HQ, represented for each patient. Based on the working hypothesis of persistent virus underlying long COVID, this triplet was thought to best represent long COVID risk attributable to this mechanism.

The ROC analysis was performed by systematically changing the thresholds in the three nested groups (in order) AC, HQ concentration and HQ%; a positive long COVID classification was obtained using Boolean logic if any of AC, HQ or HQ% were below the threshold in each of the nest loops. The sensitivity and specificity at each nested threshold point was then calculated and plotted in a ROC plot. The optimum cut-off point was determined in two ways from the ROC curve: the Youden’s J statistic and Euclidean or Upper Left Index. The area under the curve (AUC) of the multi-ROC analysis is calculated from the maximum, mean, and minimum sensitivity at each possible specificity value and calculated with numerical integration.

A series of variations of this analysis were performed: with and without the outlier exclusion, using an IQR based exclusions method, and using mean and median normalised results, where all concentration and high-quality concentration results were divided by the relevant average of the CC cohort in that variant channel. The logic of this approach was to equally weight antibody deficiencies towards each variant. A full table of results can be found in Table 3.

A more complex ROC analysis was also tested using median-normalised data, whereby data from all channels was combined for each sample for each metric. For each threshold triplet (AC, HQ, HQ%) the sample was given a ‘Vulnerability Index’ score, equal to the number of channels that fell below the thresholds being tested. A ROC was used to find the optimum cut-off value of the Vulnerability Index to discriminate positive and negative samples.

## Results

An antibody spectrum for quantity (AC), High Quality (HQ) and HQ% was recorded for each patient in all three cohorts and example spectra are shown in Figure 1: (A-C) an example of a CC patient who has developed good quality antibodies to all variants; (D-F) a patient with poor antibody quantity and inconsistent quality; and (G-I) a patient with extraordinarily high concentrations of antibodies to the early variants, all of exceptionally high quality.

**Figure 1.**
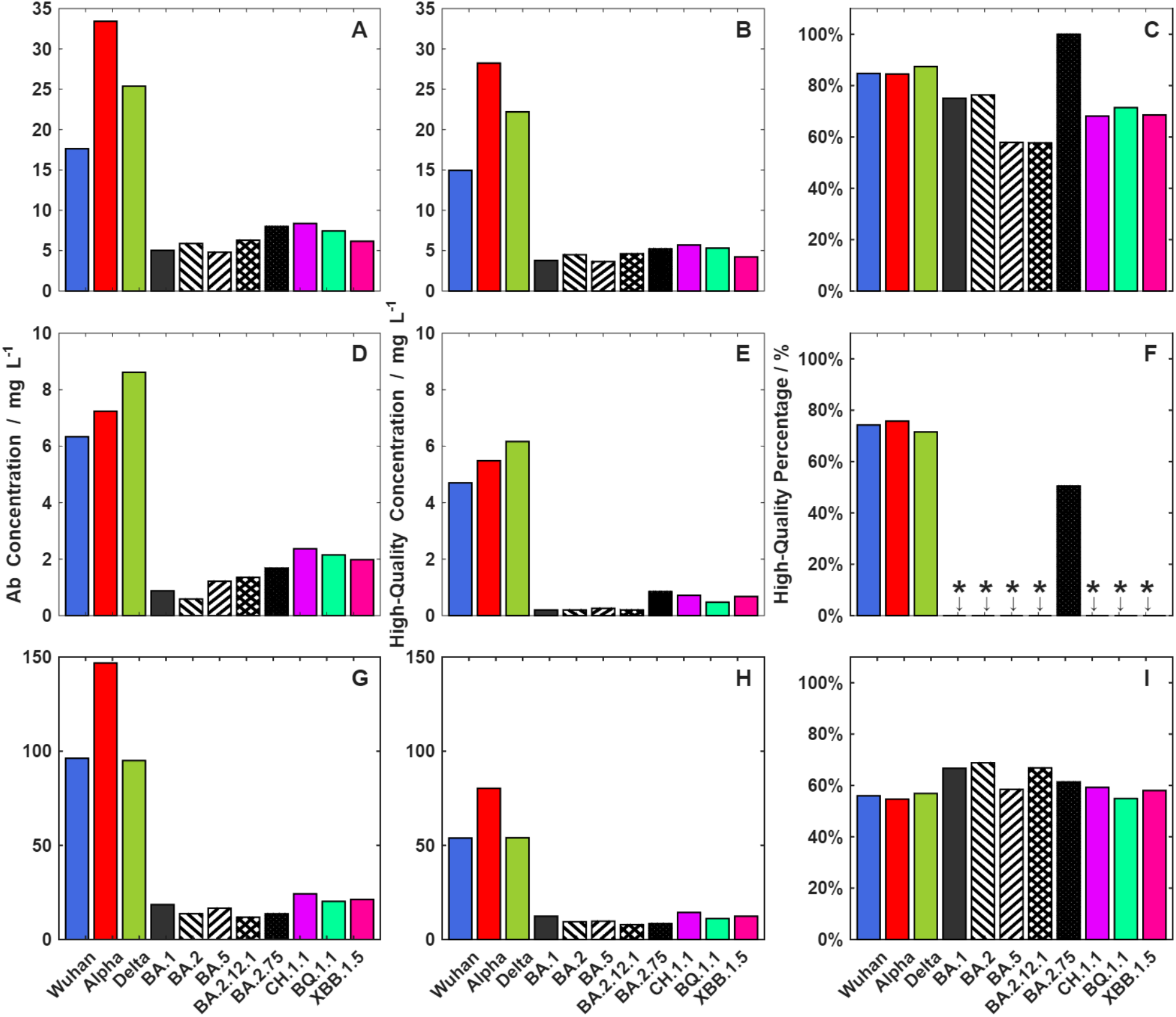
Representative Antibody Spectra: (A-C) a control patient; (D-F) a LC(+) patient and (G-I) a patient showing high antibody concentrations.

The distributions of AC, HQ and HQ% as a function of genetic distance^16^ of all variants from the escape Wuhan virus are shown in Figure 2. The AC and HQ concentrations are significantly higher (P<0.05 Mann Whitney-U) for the earlier variants compared with the later, post-omicron variants. The HQ% however remains constant: AC and HQ are correlated (*R*^2^ = 0.85 ±0.02, *m* = 0.60 ± 0.03) but AC and HQ% (*R*^2^ = 0.02 ± 0.00, *m* = 0.08 ± 0.10), and HQ and HQ% (*R*^2^ = 0.04 ± 0.05, *m* = 0.52 ± 0.04), are less so, Table S1. All three are considered variables in the subsequent analysis.

**Figure 2.**
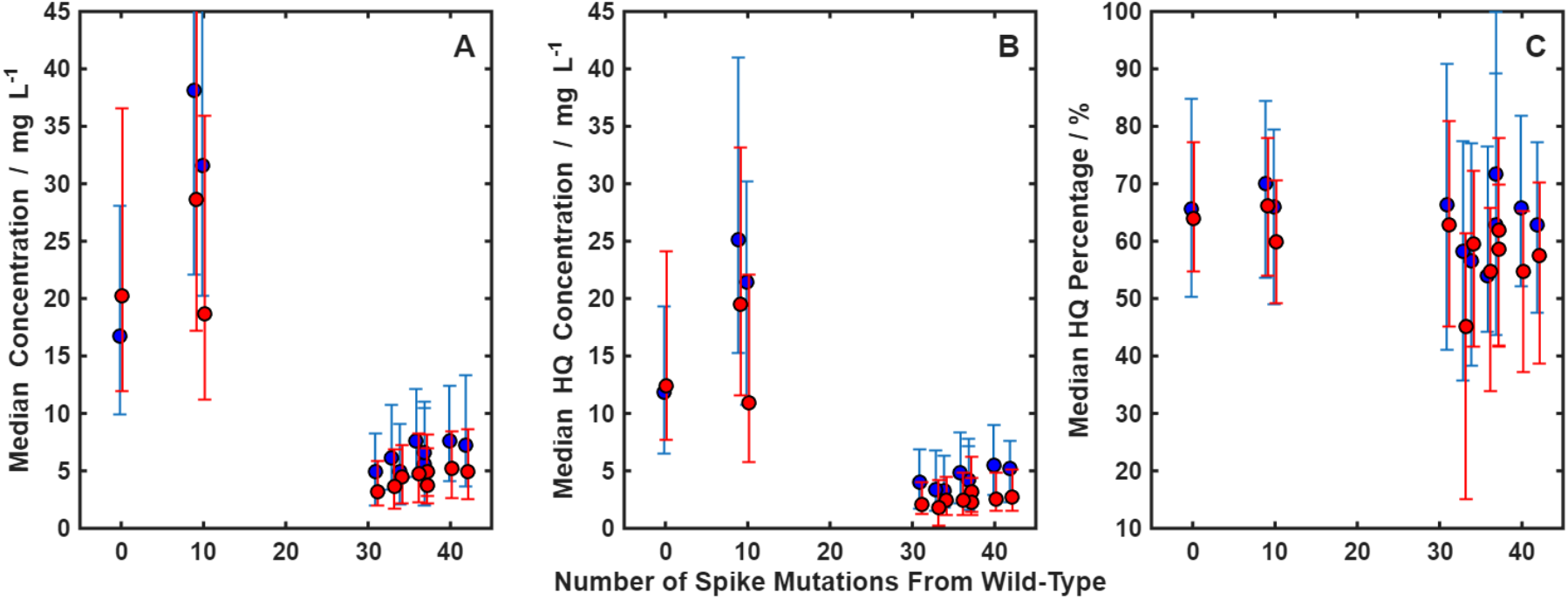
Median and interquartile range plots as a function of genetic distance from the Wuhan variant showing (A) antibody concentration (AC); (B) high-quality concentration (HQ) antibodies and (C) the percentage of high-quality antibodies (HQ%). All antibody concentrations are significantly higher for the earlier variants (P< 0.05, Mann-Whitney-U) although the proportion of high-quality antibodies (HQ%) is mostly not significantly different. Blue shows the CC cohort and red the LCC.

**Figure 3.**
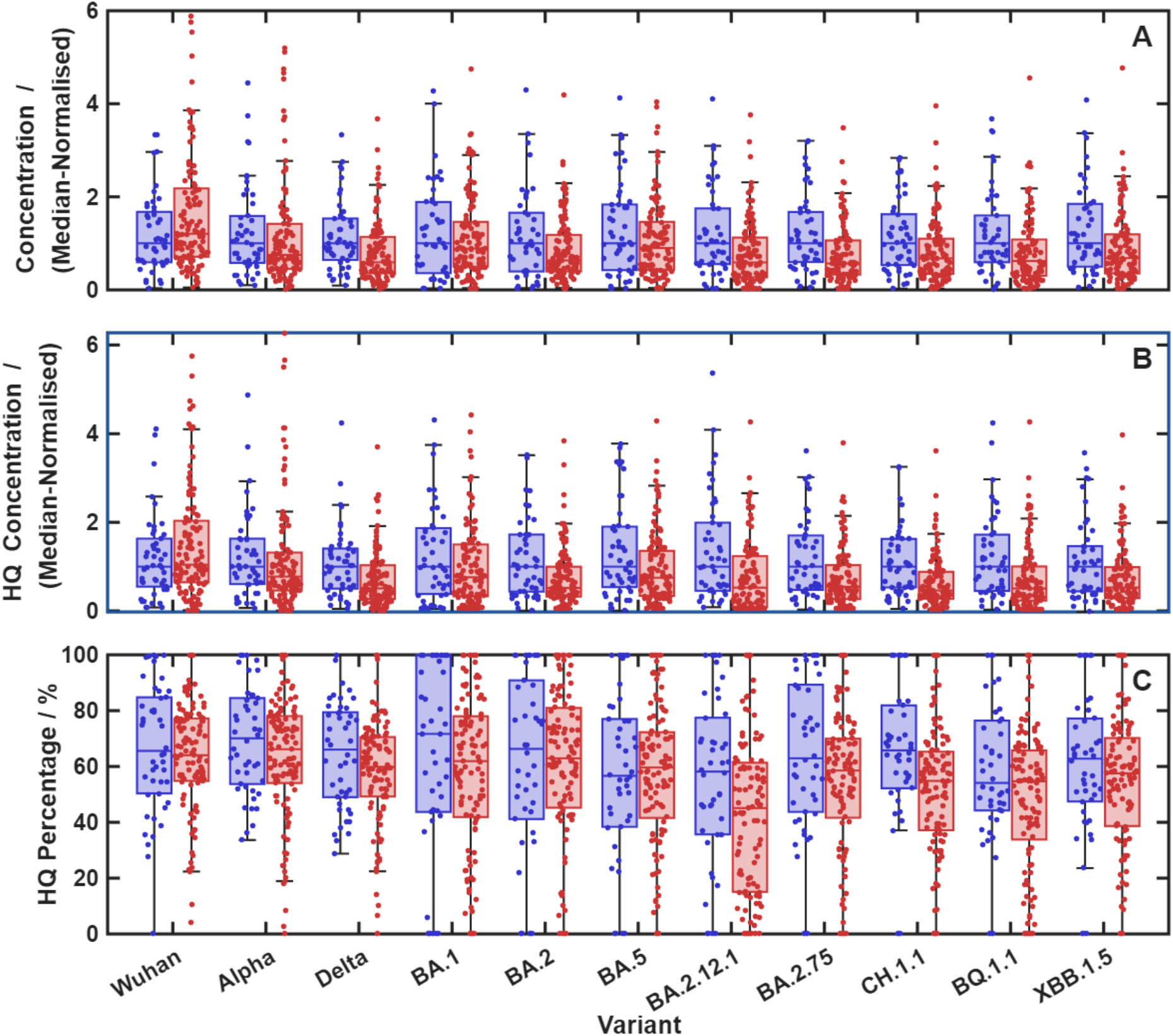
CC (blue) and LCC (red) distributions for A) AC, B) HQ and C) HQ% measures. Boxplots show medians and IQR, with the whiskers falling at the outermost non-outlier point. Outliers are any that are more than 1.5xIQR away from the median.

An outlier classification was based on the one-sided Z score = 2, with the indicted values, in mg/L, for the three pre-omicron variants then being averaged to give a single threshold value above which samples would be classified as ‘hyper’ if any variant exceeded it. The larger LCC cohort produces the following incidence: persistent virus 62% (95% CI 52% – 71%), hyperimmune 12% (95% CI 7% - 20%) and unclassified, 26% (95% CI 18% - 35%).

The nested-ROC triple-curve analysis for the CC vs LCC cohort is shown in Figure 4: the bee-swarm plot for the persistent variant risk (minimum variant HQ) selected from the spectra from each of the patients is shown with the thresholds derived from the ROC analysis. The ROC curve for the 137 patients in the CC and LCC cohorts not excluded by outlier analysis is shown in Figure 4(d) with the (coincident) Youden and Euclidean distances indicated. The summary sensitivity and specificity data and test accuracy for the ROC thresholds are collected in Table 2 and indicate a better sensitivity and specificity for the CC and LCC cohorts: 71% and 66% by Youden and Euclidean metrics.

**Table 2.** ROC thresholds and resulting diagnostic classification for CC *vs* LCC cohort after exclusion of samples falling above the Z=2 value. The full collection of results from different analysis methods can be found in Table 3.

|  | Antibody<br>Concentration<br>(mg/L) | HQ<br>Concentration<br>(mg/L) | HQ<br>Percentage<br>(%) | AUC | Sensitivity | Specificity |
| --- | --- | --- | --- | --- | --- | --- |
| CC vs<br>LCC | 0.8 | 1.5 | 34 | 0.61 (0.53-0.70) | 71 | 66 |

**Table 3.** The results from a series of ROC analyses of the CC and LCC cohorts. Where Sensitivity/Specificity/Accuracy values are provided in brackets () they include the results of the outlier analysis in the overall confusion matrix.

| **Method** | **Outlier Analysis Result** | **Sens** | **Spec** | **Accuracy** | **AUC** | **Comments** |
| --- | --- | --- | --- | --- | --- | --- |
| **Minimum HA Channel – Zcut (Z=2)** | 84.9 mg/L<br>8 CC cut, 14 LCC cut | 0.707<br>(0.743) | 0.658<br>(0.543) | 0.693<br>(0.686) | 0.614<br>(0.527-0.701) | Thresholds = 0.78/1.54/33.8 |
| **Minimum HA Channel – Zcut (Z=3.5)** | 122.5 mg/L<br>2 CC cut, 9 LCC cut | 0.707<br>(0.664) | 0.658<br>(0.652) | 0.693<br>(0.660) | 0.588<br>(0.495-0.680) | Thresholds = 0.21/1.54/0 |
| **Minimum HA Channel – No Zcut** | - | 0.699 | 0.609 | 0.673 | 0.589<br>(0.502-0.676) | Thresholds = 0.21/1.95/0 |
| **Mean-Normalised, Minimum HA Channel – Zcut (Z=2)** | 2.4 normalised mg/L<br>11 CC cut, 19 LCC cut | 0.713<br>(0.761) | 0.571<br>(0.435) | 0.674<br>(0.667) | 0.573<br>(0.472 – 0.673) | Thresholds = 0.029/0.318/0 |
| **Mean-Normalised, Minimum HA Channel – No Zcut** | - | 0.743 | 0.587 | 0.698 | 0.594<br>(0.500-0.688) | Thresholds = 0.029/0.411/0 |
| **Median-Normalised, Minimum HA Channel – Zcut (Z=2)** | 2.9 normalised mg/L<br>12 CC, 20 LCC | 0.742<br>(0.788) | 0.559<br>(0.413) | 0.693<br>(0.679) | 0.591<br>(0.400 – 0.693) | Thresholds = 0.157/0.378/0 |
| **Median-Normalised, Minimum HA Channel – No Zcut** | - | 0.726 | 0.614 | 0.694 | 0.618<br>(0.525 – 0.711) | Thresholds = 0.157/0.454/0 |
| **Median-Normalised, Minimum HA Channel Q3+1.5\*IQR cut** | 9 CC, 17 LCC | 0.823<br>(0.850) | 0.541<br>(0.435) | 0.744<br>(0.730) | 0.603<br>(0.503 – 0.702) | Thresholds = 0.157/0.484/0 |
| **Median Normalised All channels ROC – Zcut (Z=2)** | 2.9 normalised mg/L<br>11 CC, 20 LCC | 0.828 | 0.515 | 0.746 | 0.636<br>(0.559 – 0.713) | ‘Vulnerability Index’ – see Methods |

**Figure 4.**
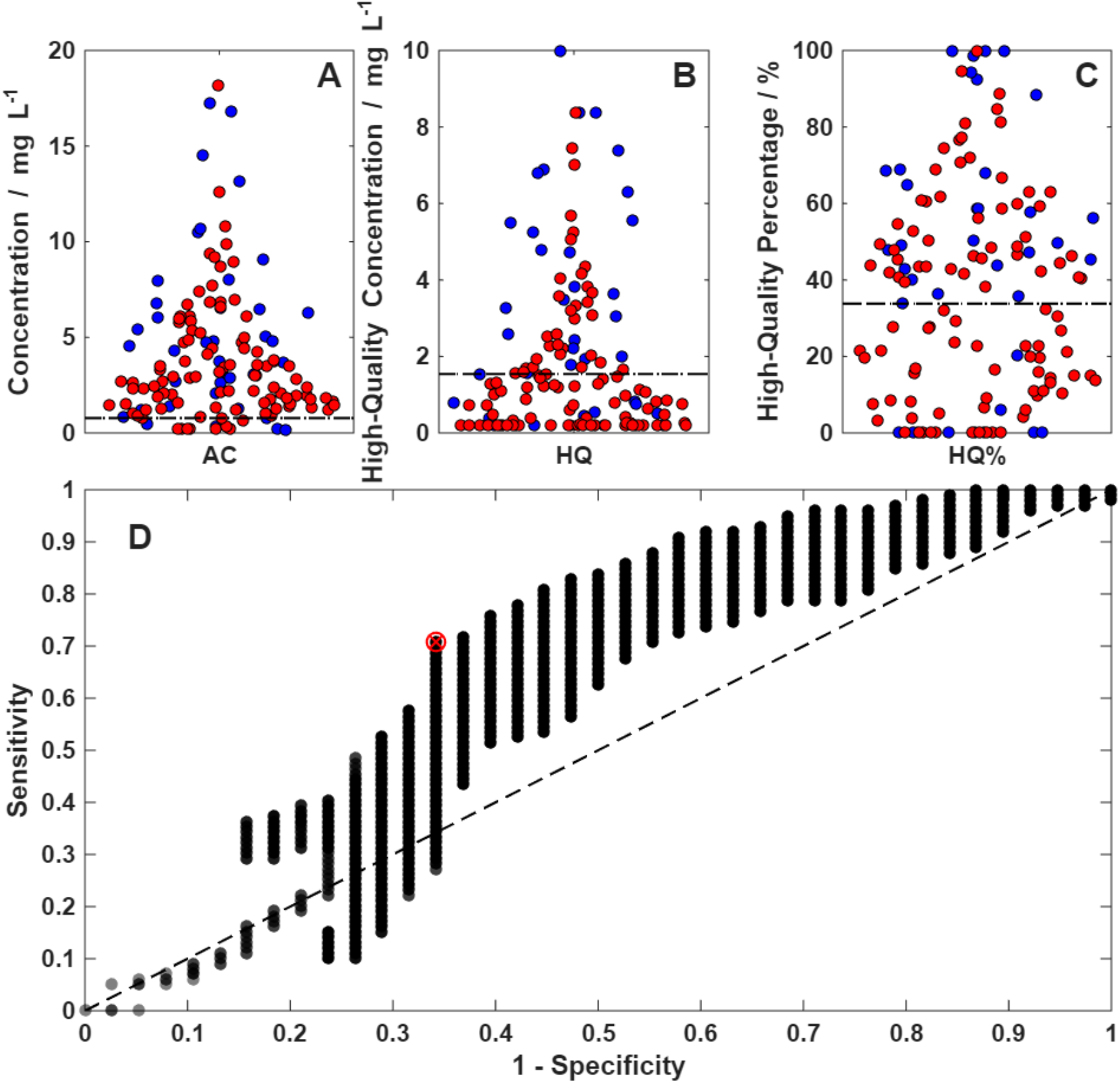
Bee Swarm Box plots for the lowest concentration high-quality antibodies: (A) antibody concentration (AC); (B) High quality antibodies (HQ); (C) percentage HQ antibodies and (D) the resulting ROC curve with the coincident Youden and Euclidean Indices marked; AUC=0.61 (0.53-0.70).

A hyperimmune cohort classification may be derived from the CC cohort and is based the outliers above the 97^th^ percentile (one-sided Z-score = 2) and gives a threshold per variant AC. The threshold applied is the mean of the three pre-omicron variants, as AC values in these channels are significantly higher than post-omicron values. The total classification in the LCC cohort is: persistent virus 62% (95% CI), hyperimmune 12% (95% CI) and unclassified, 26% (95% CI).

## Discussion

Developing a diagnostic accuracy study for a long COVID test presents significant challenges given the diverse subset of a possible 250 symptoms^1^ and the lack of a ‘gold standard’ dichotomous marker. The difficulty is reflected in the bias in cohort selection. The CC and LC cohorts were selected according to patient-reported symptom persistence recommended by the NHS with 12-week symptoms post sequalae, not the WHO^2^ 20-week persistence. It is worth noting one human disease-symptom map^17^ of 4000 human diseases shows only 400 symptoms, 250 of which have been associated with long COVID. The LCC cohort is self-selected and reviewed by the private clinician; information relating to the range of symptoms, specific ages and co-morbidities are not collected, with no limitation on symptoms lasting 12-weeks or more.

Similar problems exist for the self-classifying of control samples. Healthy controls may report no long COVID symptoms, not having associated any autoimmune disease symptoms with high antibody levels to the spike protein. Further, with asymptomatic infections, elevated antibody levels may occur which wane over time and are transiently hyperimmune.

The ROC analysis and Z=2 outlier analysis for the CC *vs* LCC cohorts leads to a robust classification of Long COVID into three types: hypo-immune or immuno-compromised, 62%, hyperimmune 12% and antibody normal, 26%. The combined diagnostic accuracy of the classification is 69% for the LCC cohort. The analysis maps directly to the pathophysiologies of persistent virus or hypo-immune, and autoimmune or hyperimmune. The triple-nested ROC analysis generated three thresholds; the antibody concentration of 0.8 mg/L, the concentration of high-quality antibodies of 1.5 mg/L, of which more than 34% need to be of high-quality to lead to clearance of the virus from the body. Similar serum characteristics have been derived for patients recovering from infection and clearing the virus^18^ with a total antibody concentration of 1.8 mg/L and the mean percentage of high-quality antibodies in a healthy cohort^19^ of 49% with (IQR 32% – 68%). The consequences for viral clearance and persistence suggests there is now a clear window post infection, during which clearance must be completed before antibody levels in the blood fall below 1.5 mg/L (reflecting partitioning throughout the body compartments) and the antibody needing to be of high-quality, corresponding with virus-antibody having a half-life of greater than 2 hours, in order to affect clearance by phagocytosis or similar mechanisms.

The population-level distributions, Figure 2, are significantly higher (Bonferroni adjusted Mann-Whitney-U, P< 0.05) for the early variants compared with the later variations. Interestingly, the median HQ% indicates that although the concentrations are low, the clonal maturation process is constant with genetic distance. The plot of antibody level *vs* genetic distance appears to show the concept of original antigenic sin ^21-23^ favouring the production of early variant antibodies, although we have established elsewhere this can be broken^19^. The trend with genetic distance may reflect the distribution of epitopes conserved in the mature clones of the population. As mutations accumulate, the number of effective clones decreases, potentially indicating only a small number of conserved epitopes, and thus a risk of vaccine evasion.

The classification of long COVID patients into three groups points to possible interventions. Patients with hypo-immune responses, suggesting persistence virus, would benefit from antibody infusions of monoclonal antibodies targeting the early variants, particularly when correlated with gaps in their antibody spectra. Some successful interventions have been reported to reduce viral load^24^; with active infections and recovery from long COVID in four immunocompromised patients^25^. Hyperimmune patients may not respond well to these monoclonal antibody interventions but better to immune suppression. Various autoimmune antibodies have been proposed to target the ACE2 receptor itself^9,26,27^ The other, ‘normal’ antibody cohort needs further differential diagnosis for other chronic conditions which may be contributing to symptoms, such as Lyme Disease, ME/CFS or mould.

## Conclusions

The mass-standardised antibody variant spectrum for antibody quantity and quality offers new insight into the persistent virus and hyperimmune mechanisms underlying the symptoms seen in long COVID. However, it is clear that all interventional studies based on symptoms alone are treating a heterogenous cohort of patients that are immune-compromised, 62%, hyperimmune, 12%, or unknown, 26%. The mass-standardised technique may be extended to CFS+/ME and the Epstein Bar virus antibody spectrum, along with the bacterial antigens responsible for Lyme or Long Lyme.

## Data Availability

All data produced in the present study are available upon reasonable request to the authors

## Funding

The project was funded by Attomarker Ltd, the alumni of the University of Exeter donating to the Covid appeal in Prof Shaw’s group, the patients taking the research test privately and Re:Cognition Health Clinic. Attomarker Ltd funded a PhD studentship for Philip H. James-Pemberton at the University of Exeter and Attomarker Ltd directly. Poppy J. Wagerfield, Ciara E. Watson, Luca Hervada worked on the study as part of their undergraduate studies at the University of Exeter.

## Declaration of Interest

Prof Shaw is the Founder, CEO and Director of Attomarker Ltd, a spin-out company from his research group at the University of Exeter

## Supplementary Data

**Table S1.** The correlation between AC, HQ and HQ%, split by cohort.

| **AC vs HQ** | **m** | **c** | **R<sup>2</sup>** |
| --- | --- | --- | --- |
| **Controls** | 0.621 | 0.678 | 0.865 |
| **LCC** | 0.578 | 0.566 | 0.840 |
| **AC vs HQ%** |  |  |  |
| **Controls** | 0.0167 | 59.8 | 0.015 |
| **LCC** | 0.151 | 53.3 | 0.018 |
| **HQ vs HQ%** |  |  |  |
| **Controls** | 0.544 | 56.8 | 0.007 |
| **LCC** | 0.490 | 51.3 | 0.073 |

## Significance tables

### Within-Group Mann Whitney U tests (Bonferroni Adjusted)

**Table S2.** Significance tables presenting the within-group Mann-Whitney U test results (Bonferroni adjusted for multiple comparisons).

| CC / AC | Wuhan | Alpha | Delta | BA.1 | BA.2 | BA.5 | BA.2.12.1 | BA.2.75 | CH.1.1 | BQ.1.1 | XBB.1.5 |
| --- | --- | --- | --- | --- | --- | --- | --- | --- | --- | --- | --- |
| Wuhan | 1.00 | 0.00 | 0.01 | 0.00 | 0.00 | 0.00 | 0.00 | 0.00 | 0.00 | 0.00 | 0.00 |
| Alpha | 0.00 | 1.00 | 1.00 | 0.00 | 0.00 | 0.00 | 0.00 | 0.00 | 0.00 | 0.00 | 0.00 |
| Delta | 0.01 | 1.00 | 1.00 | 0.00 | 0.00 | 0.00 | 0.00 | 0.00 | 0.00 | 0.00 | 0.00 |
| BA.1 | 0.00 | 0.00 | 0.00 | 1.00 | 1.00 | 1.00 | 1.00 | 1.00 | 1.00 | 0.78 | 0.62 |
| BA.2 | 0.00 | 0.00 | 0.00 | 1.00 | 1.00 | 1.00 | 1.00 | 0.85 | 0.26 | 0.13 | 0.12 |
| BA.5 | 0.00 | 0.00 | 0.00 | 1.00 | 1.00 | 1.00 | 1.00 | 1.00 | 0.80 | 0.35 | 0.38 |
| BA.2.12.1 | 0.00 | 0.00 | 0.00 | 1.00 | 1.00 | 1.00 | 1.00 | 1.00 | 1.00 | 1.00 | 1.00 |
| BA.2.75 | 0.00 | 0.00 | 0.00 | 1.00 | 0.85 | 1.00 | 1.00 | 1.00 | 1.00 | 1.00 | 1.00 |
| CH.1.1 | 0.00 | 0.00 | 0.00 | 1.00 | 0.26 | 0.80 | 1.00 | 1.00 | 1.00 | 1.00 | 1.00 |
| BQ.1.1 | 0.00 | 0.00 | 0.00 | 0.78 | 0.13 | 0.35 | 1.00 | 1.00 | 1.00 | 1.00 | 1.00 |
| XBB.1.5 | 0.00 | 0.00 | 0.00 | 0.62 | 0.12 | 0.38 | 1.00 | 1.00 | 1.00 | 1.00 | 1.00 |

| CC / HQ | Wuhan | Alpha | Delta | BA.1 | BA.2 | BA.5 | BA.2.12.1 | BA.2.75 | CH.1.1 | BQ.1.1 | XBB.1.5 |
| --- | --- | --- | --- | --- | --- | --- | --- | --- | --- | --- | --- |
| Wuhan | 1.00 | 0.00 | 0.03 | 0.00 | 0.00 | 0.00 | 0.00 | 0.00 | 0.00 | 0.00 | 0.00 |
| Alpha | 0.00 | 1.00 | 1.00 | 0.00 | 0.00 | 0.00 | 0.00 | 0.00 | 0.00 | 0.00 | 0.00 |
| Delta | 0.03 | 1.00 | 1.00 | 0.00 | 0.00 | 0.00 | 0.00 | 0.00 | 0.00 | 0.00 | 0.00 |
| BA.1 | 0.00 | 0.00 | 0.00 | 1.00 | 1.00 | 1.00 | 1.00 | 1.00 | 1.00 | 1.00 | 1.00 |
| BA.2 | 0.00 | 0.00 | 0.00 | 1.00 | 1.00 | 1.00 | 1.00 | 1.00 | 1.00 | 1.00 | 1.00 |
| BA.5 | 0.00 | 0.00 | 0.00 | 1.00 | 1.00 | 1.00 | 1.00 | 1.00 | 0.45 | 1.00 | 1.00 |
| BA.2.12.1 | 0.00 | 0.00 | 0.00 | 1.00 | 1.00 | 1.00 | 1.00 | 1.00 | 0.72 | 1.00 | 1.00 |
| BA.2.75 | 0.00 | 0.00 | 0.00 | 1.00 | 1.00 | 1.00 | 1.00 | 1.00 | 1.00 | 1.00 | 1.00 |
| CH.1.1 | 0.00 | 0.00 | 0.00 | 1.00 | 1.00 | 0.45 | 0.72 | 1.00 | 1.00 | 1.00 | 1.00 |
| BQ.1.1 | 0.00 | 0.00 | 0.00 | 1.00 | 1.00 | 1.00 | 1.00 | 1.00 | 1.00 | 1.00 | 1.00 |
| XBB.1.5 | 0.00 | 0.00 | 0.00 | 1.00 | 1.00 | 1.00 | 1.00 | 1.00 | 1.00 | 1.00 | 1.00 |

| CC / HQ% | Wuhan | Alpha | Delta | BA.1 | BA.2 | BA.5 | BA.2.12.1 | BA.2.75 | CH.1.1 | BQ.1.1 | XBB.1.5 |
| --- | --- | --- | --- | --- | --- | --- | --- | --- | --- | --- | --- |
| Wuhan | 1.00 | 1.00 | 1.00 | 1.00 | 1.00 | 1.00 | 0.97 | 1.00 | 1.00 | 1.00 | 1.00 |
| Alpha | 1.00 | 1.00 | 1.00 | 1.00 | 1.00 | 0.47 | 0.26 | 1.00 | 1.00 | 0.22 | 1.00 |
| Delta | 1.00 | 1.00 | 1.00 | 1.00 | 1.00 | 1.00 | 1.00 | 1.00 | 1.00 | 1.00 | 1.00 |
| BA.1 | 1.00 | 1.00 | 1.00 | 1.00 | 1.00 | 1.00 | 1.00 | 1.00 | 1.00 | 1.00 | 1.00 |
| BA.2 | 1.00 | 1.00 | 1.00 | 1.00 | 1.00 | 1.00 | 1.00 | 1.00 | 1.00 | 1.00 | 1.00 |
| BA.5 | 1.00 | 0.47 | 1.00 | 1.00 | 1.00 | 1.00 | 1.00 | 1.00 | 1.00 | 1.00 | 1.00 |
| BA.2.12.1 | 0.97 | 0.26 | 1.00 | 1.00 | 1.00 | 1.00 | 1.00 | 1.00 | 0.69 | 1.00 | 1.00 |
| BA.2.75 | 1.00 | 1.00 | 1.00 | 1.00 | 1.00 | 1.00 | 1.00 | 1.00 | 1.00 | 1.00 | 1.00 |
| CH.1.1 | 1.00 | 1.00 | 1.00 | 1.00 | 1.00 | 1.00 | 0.69 | 1.00 | 1.00 | 0.64 | 1.00 |
| BQ.1.1 | 1.00 | 0.22 | 1.00 | 1.00 | 1.00 | 1.00 | 1.00 | 1.00 | 0.64 | 1.00 | 1.00 |
| XBB.1.5 | 1.00 | 1.00 | 1.00 | 1.00 | 1.00 | 1.00 | 1.00 | 1.00 | 1.00 | 1.00 | 1.00 |

| CC / AC | Wuhan | Alpha | Delta | BA.1 | BA.2 | BA.5 | BA.2.12.1 | BA.2.75 | CH.1.1 | BQ.1.1 | XBB.1.5 |
| --- | --- | --- | --- | --- | --- | --- | --- | --- | --- | --- | --- |
| Wuhan | 1.00 | 0.02 | 1.00 | 0.00 | 0.00 | 0.00 | 0.00 | 0.00 | 0.00 | 0.00 | 0.00 |
| Alpha | 0.02 | 1.00 | 0.00 | 0.00 | 0.00 | 0.00 | 0.00 | 0.00 | 0.00 | 0.00 | 0.00 |
| Delta | 1.00 | 0.00 | 1.00 | 0.00 | 0.00 | 0.00 | 0.00 | 0.00 | 0.00 | 0.00 | 0.00 |
| BA.1 | 0.00 | 0.00 | 0.00 | 1.00 | 0.01 | 1.00 | 0.11 | 0.48 | 1.00 | 1.00 | 1.00 |
| BA.2 | 0.00 | 0.00 | 0.00 | 0.01 | 1.00 | 0.90 | 1.00 | 1.00 | 0.02 | 0.10 | 0.02 |
| BA.5 | 0.00 | 0.00 | 0.00 | 1.00 | 0.90 | 1.00 | 1.00 | 1.00 | 1.00 | 1.00 | 1.00 |
| BA.2.12.1 | 0.00 | 0.00 | 0.00 | 0.11 | 1.00 | 1.00 | 1.00 | 1.00 | 0.13 | 0.40 | 0.09 |
| BA.2.75 | 0.00 | 0.00 | 0.00 | 0.48 | 1.00 | 1.00 | 1.00 | 1.00 | 0.65 | 1.00 | 0.53 |
| CH.1.1 | 0.00 | 0.00 | 0.00 | 1.00 | 0.02 | 1.00 | 0.13 | 0.65 | 1.00 | 1.00 | 1.00 |
| BQ.1.1 | 0.00 | 0.00 | 0.00 | 1.00 | 0.10 | 1.00 | 0.40 | 1.00 | 1.00 | 1.00 | 1.00 |
| XBB.1.5 | 0.00 | 0.00 | 0.00 | 1.00 | 0.02 | 1.00 | 0.09 | 0.53 | 1.00 | 1.00 | 1.00 |

| LCC / HQ | Wuhan | Alpha | Delta | BA.1 | BA.2 | BA.5 | BA.2.12.1 | BA.2.75 | CH.1.1 | BQ.1.1 | XBB.1.5 |
| --- | --- | --- | --- | --- | --- | --- | --- | --- | --- | --- | --- |
| Wuhan | 1.00 | 0.03 | 1.00 | 0.00 | 0.00 | 0.00 | 0.00 | 0.00 | 0.00 | 0.00 | 0.00 |
| Alpha | 0.03 | 1.00 | 0.00 | 0.00 | 0.00 | 0.00 | 0.00 | 0.00 | 0.00 | 0.00 | 0.00 |
| Delta | 1.00 | 0.00 | 1.00 | 0.00 | 0.00 | 0.00 | 0.00 | 0.00 | 0.00 | 0.00 | 0.00 |
| BA.1 | 0.00 | 0.00 | 0.00 | 1.00 | 0.18 | 1.00 | 0.00 | 0.42 | 1.00 | 0.77 | 1.00 |
| BA.2 | 0.00 | 0.00 | 0.00 | 0.18 | 1.00 | 1.00 | 0.49 | 1.00 | 0.90 | 1.00 | 0.35 |
| BA.5 | 0.00 | 0.00 | 0.00 | 1.00 | 1.00 | 1.00 | 0.06 | 1.00 | 1.00 | 1.00 | 1.00 |
| BA.2.12.1 | 0.00 | 0.00 | 0.00 | 0.00 | 0.49 | 0.06 | 1.00 | 0.31 | 0.01 | 0.24 | 0.00 |
| BA.2.75 | 0.00 | 0.00 | 0.00 | 0.42 | 1.00 | 1.00 | 0.31 | 1.00 | 1.00 | 1.00 | 0.87 |
| CH.1.1 | 0.00 | 0.00 | 0.00 | 1.00 | 0.90 | 1.00 | 0.01 | 1.00 | 1.00 | 1.00 | 1.00 |
| BQ.1.1 | 0.00 | 0.00 | 0.00 | 0.77 | 1.00 | 1.00 | 0.24 | 1.00 | 1.00 | 1.00 | 1.00 |
| XBB.1.5 | 0.00 | 0.00 | 0.00 | 1.00 | 0.35 | 1.00 | 0.00 | 0.87 | 1.00 | 1.00 | 1.00 |

| LCC / HQ% | Wuhan | Alpha | Delta | BA.1 | BA.2 | BA.5 | BA.2.12.1 | BA.2.75 | CH.1.1 | BQ.1.1 | XBB.1.5 |
| --- | --- | --- | --- | --- | --- | --- | --- | --- | --- | --- | --- |
| Wuhan | 1.00 | 1.00 | 0.33 | 1.00 | 1.00 | 0.79 | 0.00 | 0.10 | 0.00 | 0.00 | 0.03 |
| Alpha | 1.00 | 1.00 | 0.17 | 1.00 | 1.00 | 0.57 | 0.00 | 0.06 | 0.00 | 0.00 | 0.02 |
| Delta | 0.33 | 0.17 | 1.00 | 1.00 | 1.00 | 1.00 | 0.00 | 1.00 | 0.20 | 0.02 | 1.00 |
| BA.1 | 1.00 | 1.00 | 1.00 | 1.00 | 1.00 | 1.00 | 0.00 | 1.00 | 0.27 | 0.03 | 1.00 |
| BA.2 | 1.00 | 1.00 | 1.00 | 1.00 | 1.00 | 1.00 | 0.00 | 0.43 | 0.02 | 0.00 | 0.26 |
| BA.5 | 0.79 | 0.57 | 1.00 | 1.00 | 1.00 | 1.00 | 0.00 | 1.00 | 0.85 | 0.15 | 1.00 |
| BA.2.12.1 | 0.00 | 0.00 | 0.00 | 0.00 | 0.00 | 0.00 | 1.00 | 0.01 | 0.14 | 1.00 | 0.02 |
| BA.2.75 | 0.10 | 0.06 | 1.00 | 1.00 | 0.43 | 1.00 | 0.01 | 1.00 | 1.00 | 0.58 | 1.00 |
| CH.1.1 | 0.00 | 0.00 | 0.20 | 0.27 | 0.02 | 0.85 | 0.14 | 1.00 | 1.00 | 1.00 | 1.00 |
| BQ.1.1 | 0.00 | 0.00 | 0.02 | 0.03 | 0.00 | 0.15 | 1.00 | 0.58 | 1.00 | 1.00 | 1.00 |
| XBB.1.5 | 0.03 | 0.02 | 1.00 | 1.00 | 0.26 | 1.00 | 0.02 | 1.00 | 1.00 | 1.00 | 1.00 |

### Group Comparison Mann Whitney U Tests (Bonferroni Adjusted)

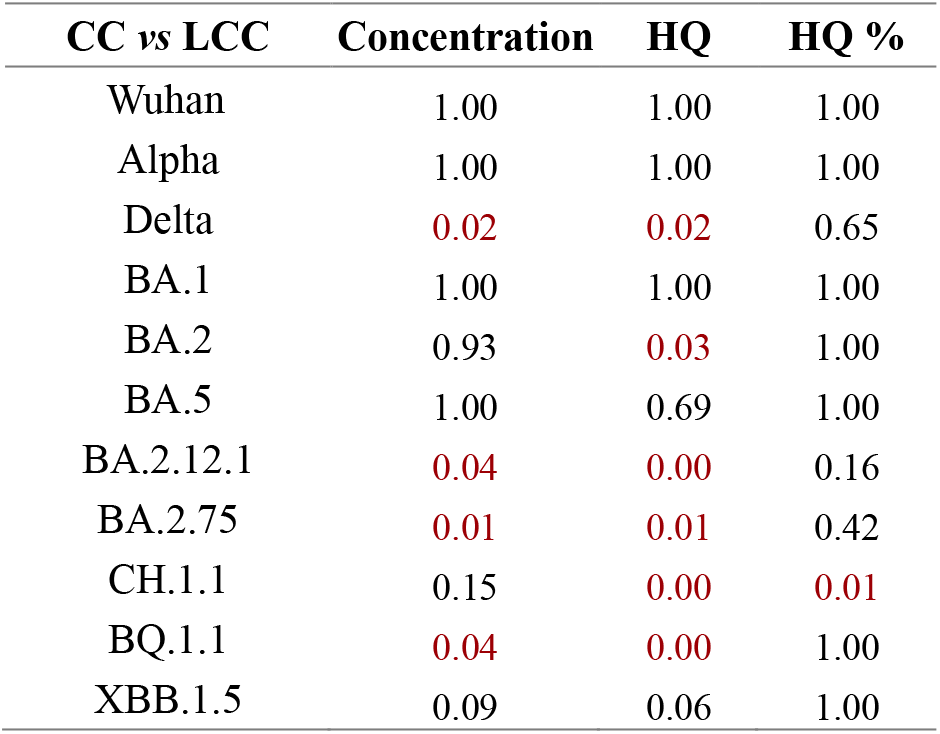

| CC vs LCC | Concentration | HQ | HQ % |
| --- | --- | --- | --- |
| Wuhan | 1.00 | 1.00 | 1.00 |
| Alpha | 1.00 | 1.00 | 1.00 |
| Delta | 0.02 | 0.02 | 0.65 |
| BA.1 | 1.00 | 1.00 | 1.00 |
| BA.2 | 0.93 | 0.03 | 1.00 |
| BA.5 | 1.00 | 0.69 | 1.00 |
| BA.2.12.1 | 0.04 | 0.00 | 0.16 |
| BA.2.75 | 0.01 | 0.01 | 0.42 |
| CH.1.1 | 0.15 | 0.00 | 0.01 |
| BQ.1.1 | 0.04 | 0.00 | 1.00 |
| XBB.1.5 | 0.09 | 0.06 | 1.00 |

## Notes

### Summary of Updates

The revision updated a broken link to a table

